# Vitamin D Status: A U-shaped relationship for SARS-CoV-2 seropositivity in UK healthcare workers

**DOI:** 10.1101/2021.10.11.21264835

**Authors:** William R Mackay, Sebastian T Lugg, Aduragbemi A Faniyi, Sian E Faustini, Craig Webster, Joanne E Duffy, Martin Hewison, Adrian M Shields, Alex G Richter, Dhruv Parekh, Aaron Scott, David R Thickett

## Abstract

**Background:** Vitamin D has numerous mechanistic roles within the immune system. There is increasing evidence to suggest Vitamin D deficiency may increase individuals’ risk of COVID-19 infection and susceptibility. We aimed to determine the relationship between severity of vitamin D deficiency and sufficiency and COVID-19 infection within healthcare workers.

**Methods:** The study included an observational cohort of healthcare workers who isolated due to COVID-19 symptoms from 12th to 22nd May 2020, from the University Hospitals Birmingham NHS Foundation Trust (UHBFT). This was part of the COVID-19 convalescent immunity study (COCO). Data collected included SARS-CoV-2 seroconversion status, serum 25(OH)D_3_ levels as well as age, body mass index (BMI), sex, ethnicity, job role, and co-morbidities. Participants were grouped into four vitamin D (VD) categories. 1) Severe VD deficiency (VD <30 nmol/L); 2) VD deficiency (30 nmol/L ≤ VD <50 nmol/L); 3) VD insufficiency (50 nmol/L ≤ VD <75 nmol/L); 4) VD sufficiency (VD ≥75 nmol/L).

**Results:** When VD levels were compared against COVID-19 seropositivity rate, a U-shaped curve was identified in the total population. This trend repeated when split into subgroups of age, sex, ethnicity, BMI, and co-morbidity status. Significant difference was identified in the COVID-19 seropositivity rate between VD groups between multiple VD groups in the total population, males, females, BAME, BMI<30 (kg/m^2^), 0 and +1 comorbidities; the majority of which were differences when the severely VD deficient category were compared to the other group. A significantly larger proportion of those within the Black, Asian, minority ethnic (BAME) group (*vs*. white ethnicity) were severely vitamin D deficient (*P <*0.00001). A significantly higher proportion of the 0-comorbidity subgroup were vitamin D deficient in comparison to the 1+ comorbidity subgroup (*P* = 0.046).

**Conclusions:** Further investigation of the U-shaped curves is required to determine whether high VD levels can have a detrimental effect on susceptibility to COVID-19 infection. Future randomised clinical trials of VD supplementation could potentially identify ‘optimal’ VD levels. This would allow for targeted therapeutic treatment for those at-risk such as in the BAME group.

## Background

Vitamin D (VD) is an essential lipophilic secosteroid that has a complex interrelationship with both the innate and adaptive immune system [1]. Numerous investigations have determined Vitamin D_3_ deficiency to drive the increase of infection [2] due to an accompanied change in functional immunity [3]. In bacterial sepsis, VD deficiency has been shown to have a role in the development of the acute respiratory distress syndrome (ARDS) development, a complication also seen in severe COVID-19 infection [4]. As an estimated 39% of COVID-19 patients with ARDS have died [5], it is vital to understand whether VD deficiency increases the risk of SARS-CoV-2 infection.

Individuals of the Black, Asian, and ethnic minority (BAME) groups appear to be disproportionately at-risk of COVID-19 [6]. VD deficiency is more prevalent in darker-skinned individuals in comparison to those of lighter skin [7]. This may in part provide some explanation for the increases risk of COVID-19 infection in those of BAME ethnicity, and whether addressing irregular serum levels could reduce infection risk.

A recent review which collated VD’s association with both COVID-19 infection and mortality found mixed results [8]. A conclusion found overall increased risk of infection and mortality, but a large proportion of studies did not control for important confounders. There is limited data on VD levels and associated COVID-19 in healthcare workers, who were at a higher risk of developing COVID-19 during the pandemic [9]. We published a rapid research letter during the pandemic of a cross-sectional study of UK healthcare workers who isolated due to symptoms of COVID-19 [10]. Our early analysis found that BAME ethnicity are at the highest risk of VD deficiency (VD levels <30 nmol/L) and that VD deficiency was an independent risk factor for development of COVID-19 seroconversion; the biggest differences in seroconversion were seen in the BAME male group. However, the association of differing degrees of deficiency and relationship with demographics, comorbidity and COVID-19 infection were not investigated.

In an established cohort of healthcare workers who isolated due to symptoms of COVID-19, this study therefore aimed to determine in detail the relationship between severity of VD deficiency and sufficiency and COVID-19 infection.

## Methods

As part of an observational study, healthcare workers were recruited from 12th to 22nd May 2020 from the University Hospitals Birmingham NHS Foundation Trust (UHBFT). This was part of the COVID-19 convalescent immunity study (COCO) approved by London - Camden & Kings Cross Research Ethics Committee (20/HRA/1817). The inclusion criteria of this cohort studied were staff members who had symptoms suggestive of COVID-19. Demographic details were obtained including age, BMI, sex, ethnicity and co-morbidities. Blood samples were taken to the laboratory for processing to obtain serum for SARS-CoV-2 antibody and for the vitamin D assay.

Anti-SARS-CoV-2 spike glycoprotein antibodies were measured using a combined IgG, IgA, IgM ELISA antibody with 98.3% (95% CI: 96.4-99.4%) specificity and 98.6% sensitivity (95% CI: 92.6-100%) (Product code MK654, The Binding Site (TBS), Birmingham) [11]. Seroconversion was used as an immunological surrogate of prior SARS-CoV-2 infection. Vitamin D status was determined by measurement of serum 25(OH)D_3_ using mass spectrometry. Concentrations of VD were reported in nmol/L and stratified into the following categories: 1) Severe VD deficiency (VD <30 nmol/L); 2) VD deficiency (30 nmol/L ≤ VD <50 nmol/L); 3) VD insufficiency (50 nmol/L ≤ VD <75 nmol/L); 4) VD sufficiency (VD ≥75 nmol/L) [12].

Data was analysed using IBM SPSS Statistics (version 27). Continuous data was reported as mean ± standard deviation (SD) or median-interquartile range (IQR) depending on the normality of distribution. Categorical data were reported via frequency and proportion. Continuous data were assessed for normality with the Shapiro-Wilk test. When VD was stratified into groups, continuous variables were compared using either Independent Samples t-test or Mann-Whitney U test. Categorical data were assessed with Fisher’s exact test. Correlation between VD and SARS-CoV-2 seropositivity were determined by a second-order polynomial regression. Statistical significance was defined as *p*-value <0.05 in all cases.

## Results

In total, there were 379 participants. The median age of the cohort was 42.0 (IQR 30.0 – 50.0) years. Representation from the cohort included 282 females (74.4%), 274 of white ethnicity (72.3%), a median BMI of 25.9 (IQR 22.9 – 30.1) kg/m^2^ and 233 (71.5%) had no underlying comorbidities. The median VD_3_ levels of the entire cohort was 55.4 (39.2-68.8) nmol/L. A further breakdown of the participant demographics is presented in *Table 1*.

**Table 1.**
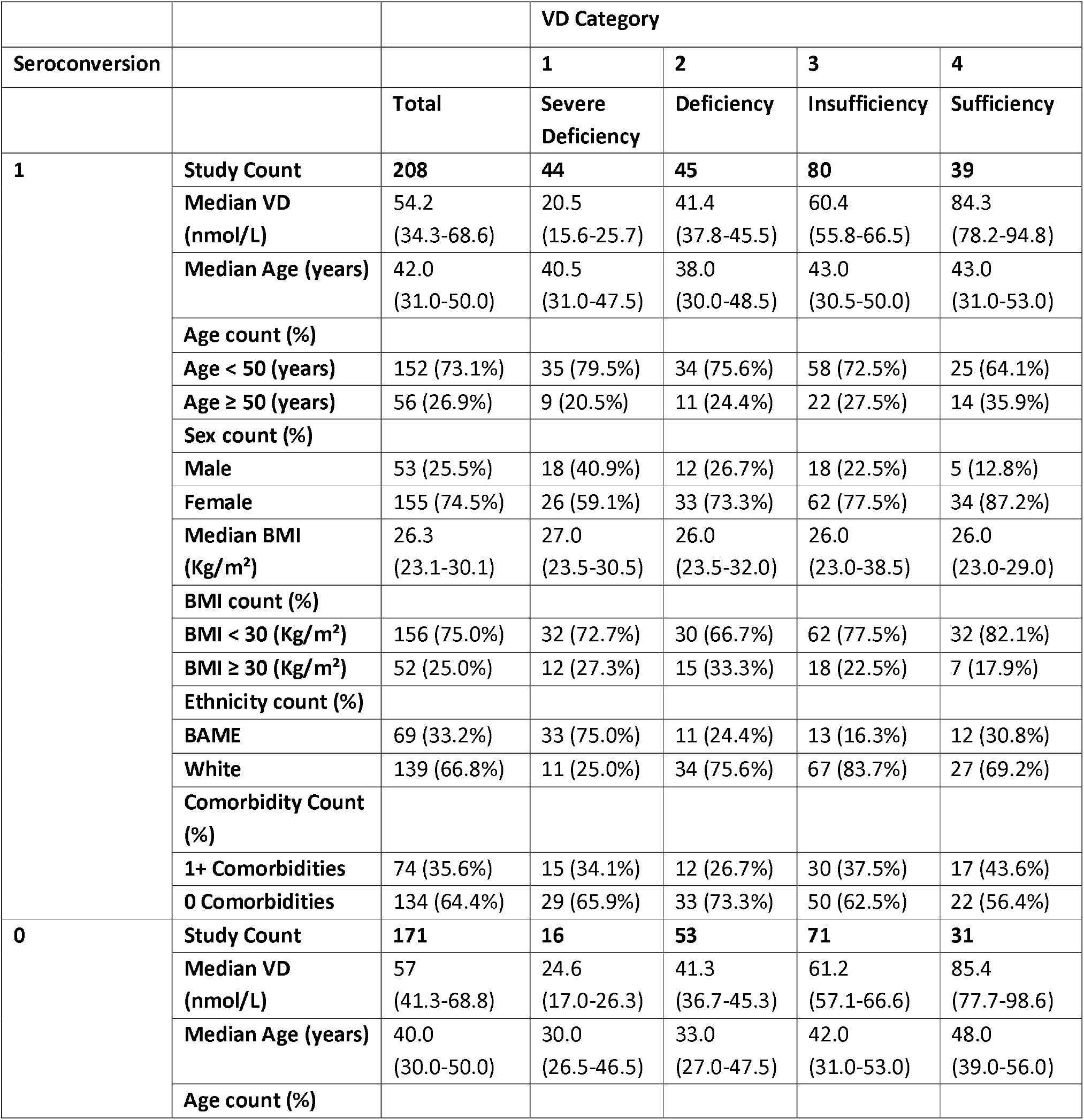
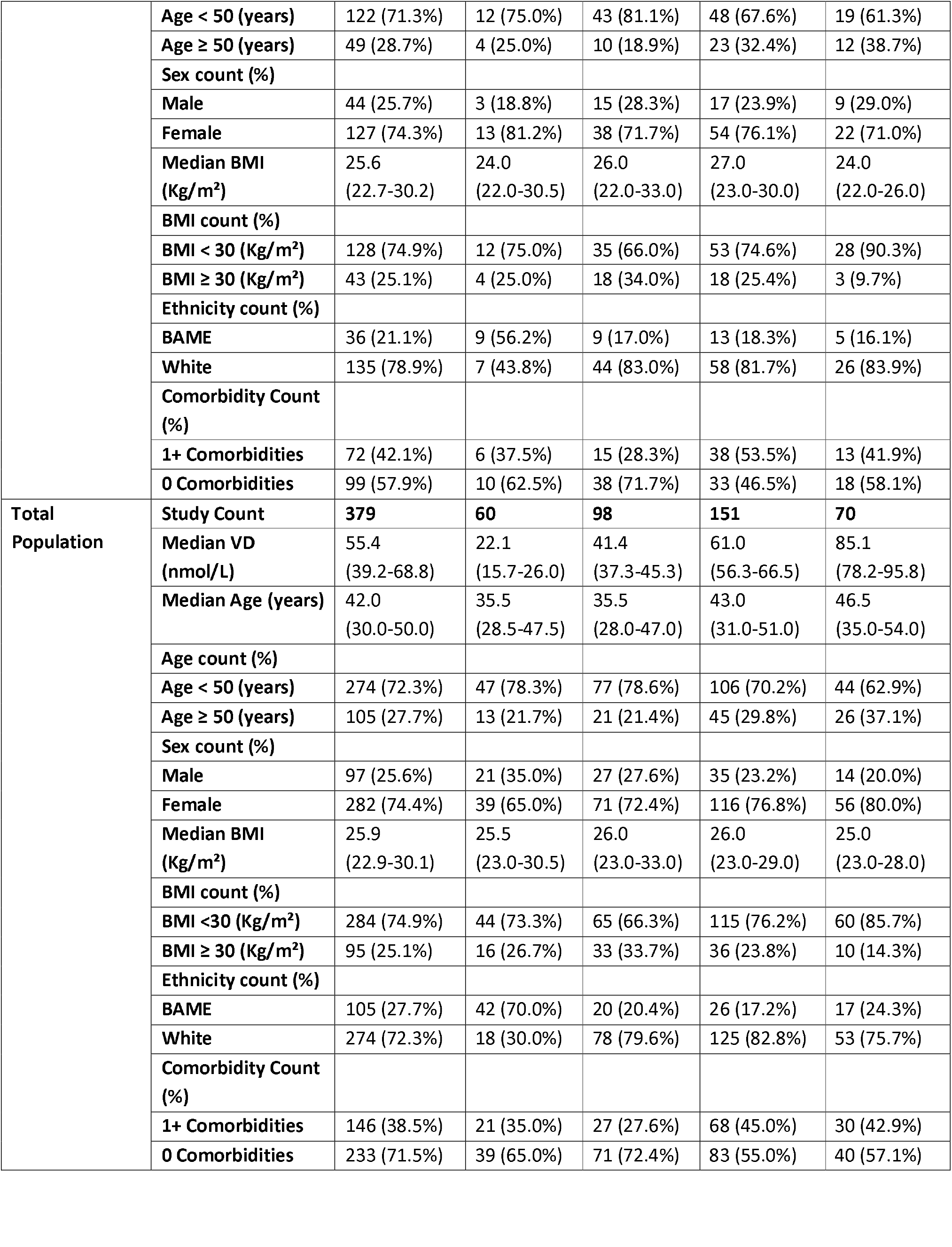
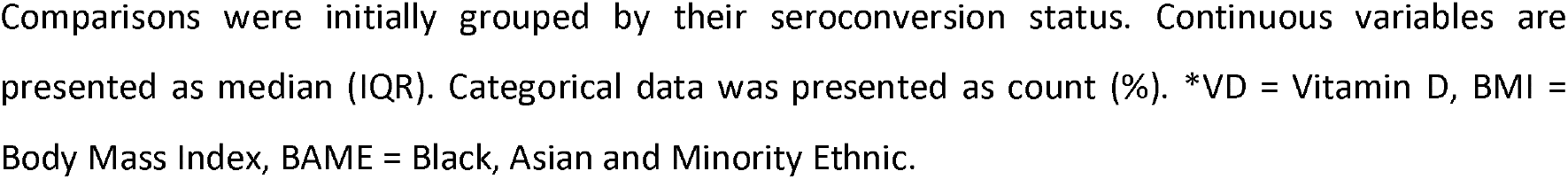
Aggregated measures within each VD category.

The total population was stratified into four VD categories, 60 (15.8%) with severe deficiency (22.1 (IQR 15.7-26.0) nmol/L), 98 (25.9%) with deficiency (IQR 41.4 (37.3-45.3) nmol/L), 151 (39.8%) with insufficiency (61.0 (IQR 56.3-66.5) nmol/L), and 70 (18.5%) with sufficiency (85.1 (IQR 78.2-95.8) nmol/L) (*Table 1*). The U-curve shown in the total population (*Figure 1A*) shows that below levels of 80 nmol/L, seropositivity increases as the VD level reduces. The trend lines for all the cohort subgroups (*Figures 1B, 1C, 1D, 1E* and *1F*) all broadly follow this U-shaped curve, with plateau points spread between 80 nmol/L and 100 nmol/L.

**Figure 1.**
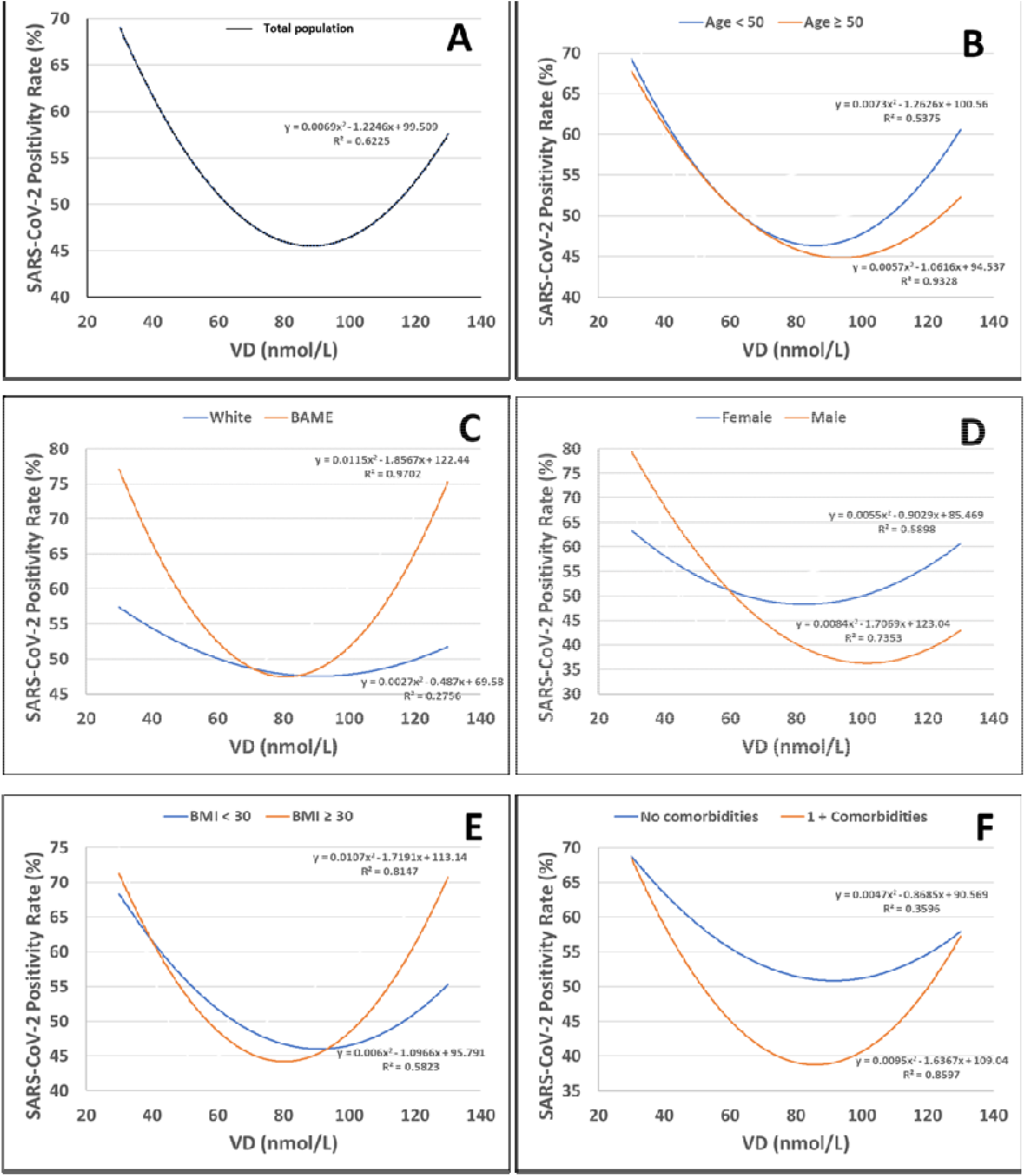
SARS-CoV-2 seropositivity rate against serum VD levels by (A) Total, (B) Age group, (C) Ethnicity, (D) Sex, (E) BMI, and (F) Presence of Comorbidities. Seropositivity rate is defined as the number of SARS-CoV-2 positive cases, divided by the total number of cases. The data is represented by a weighted second-order polynomial regression smooth line. The line equation and the R^2^ value is placed beside each corresponding line. *VD = Vitamin D, BMI = Body Mass Index, BAME = Black, Asian and Minority Ethnic.

### VD association with SARS-CoV-2 seropositivity

208 (54.9%) of the cohort tested positive for SARS-CoV-2 anti-spike glycoprotein antibodies (*Table 2*). No significant difference existed between the median VD values of the COVID-19 seropositive (54.2 (34.3-68.6) nmol/L) and COVID-19 negative group (57.0 (41.3-68.8) nmol/L) (*P* = 0.20) (*Table 1*). Inspection of the total population (*Figure 2A*) shows that the COVID-19 seropositive group had 2.75 times the amount of severely deficient participants in comparison to the COVID-19 seronegative group (*Table 2*). When VD was stratified, SARS-CoV-2 seropositivity were 73.3% in the severely deficient group compared to deficient (45.9%; *P* = 0.001), insufficient (53.0%; *P* = 0.008), and sufficient (56.7%; *P* = 0.049) (*Table 2* and *3*).

**Table 2.**
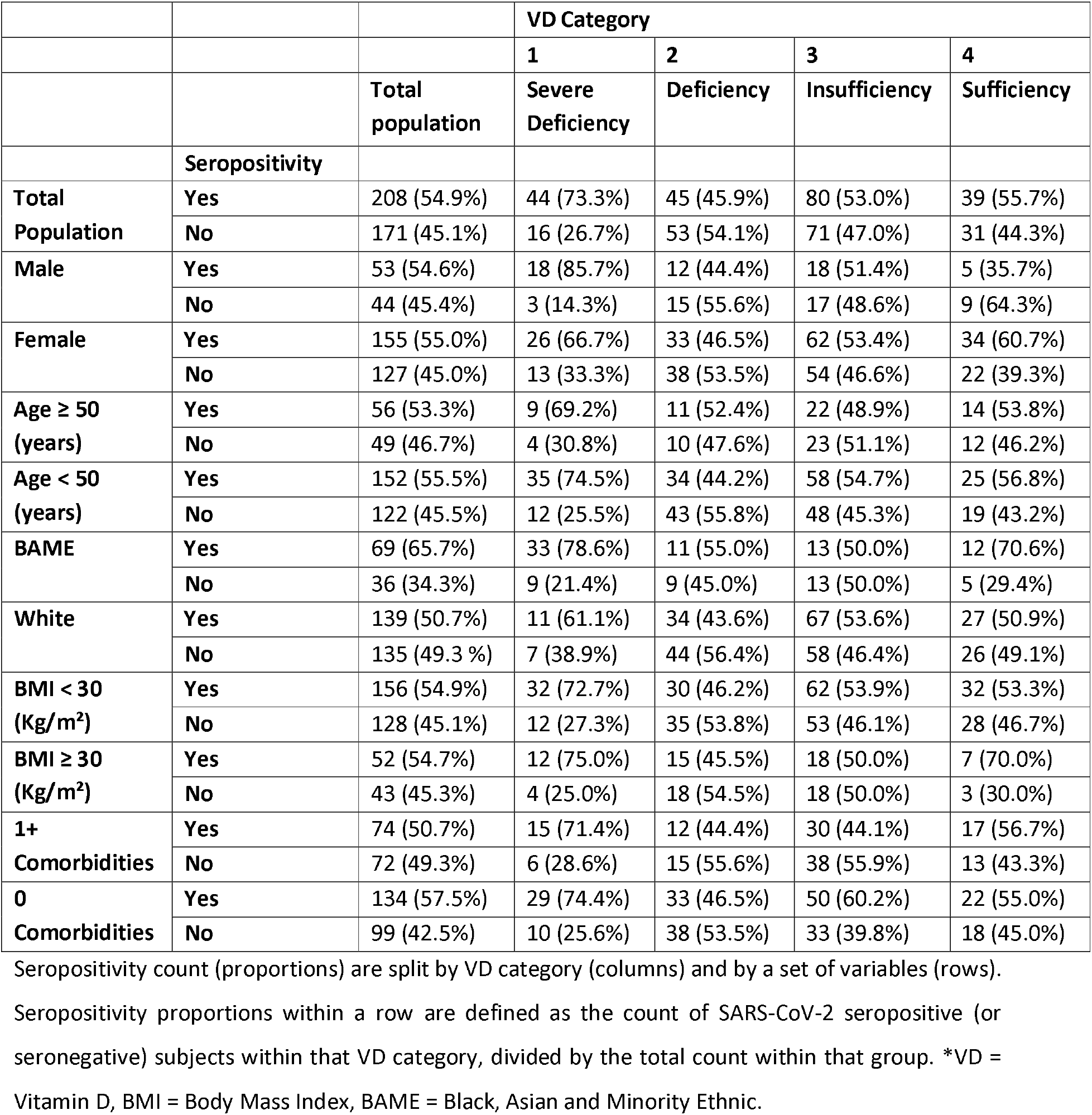
The proportion of positive and negative SARS-CoV-2 seroconversions.

**Figure 2.**
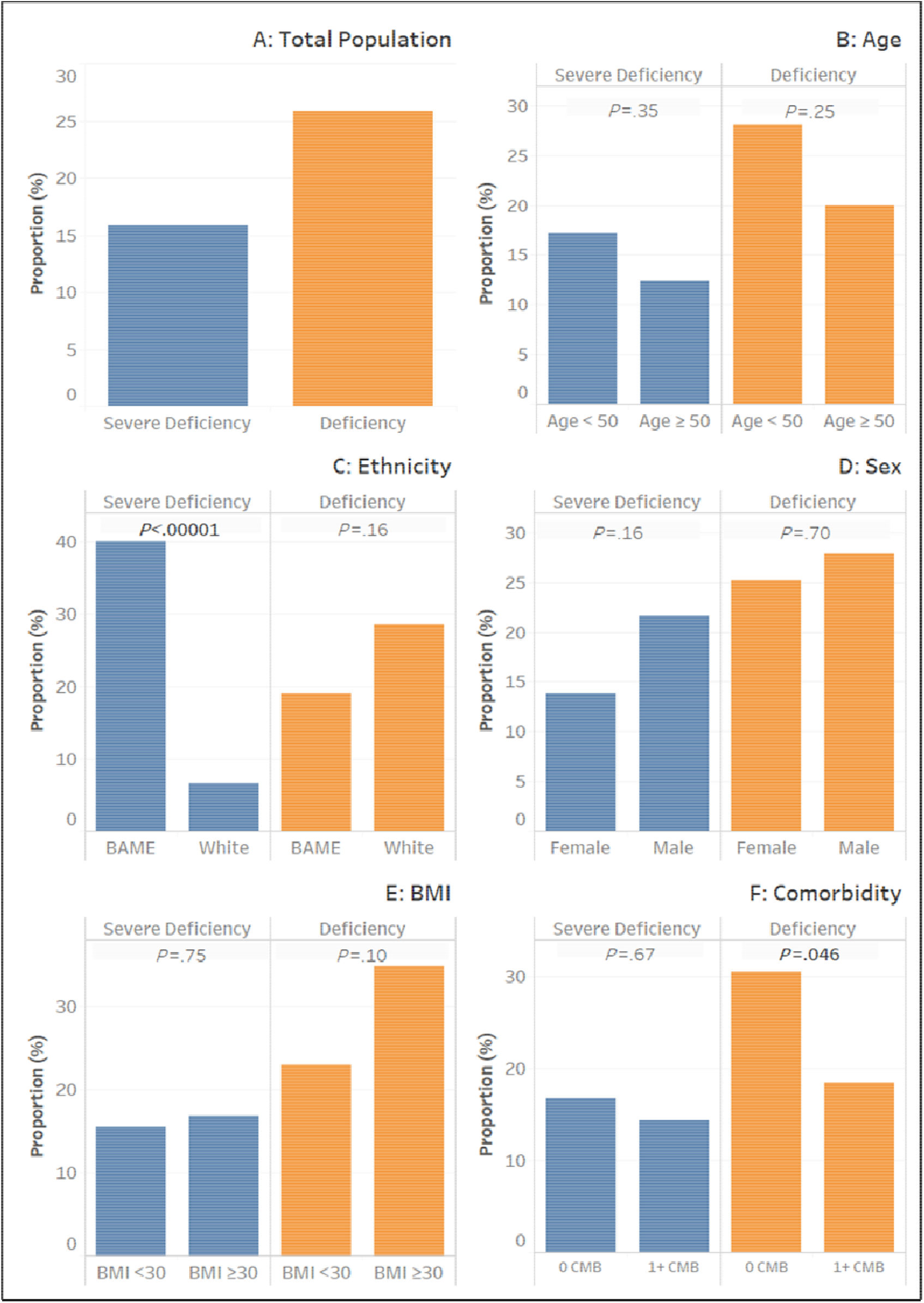
Proportion of subgroup population by VD category. Each bar represents the proportion of the specific group which are VD deficient and severely VD deficient. Comparisons are made between the two subgroups to determine whether there is a significant difference in the proportion of the subgroup which is in the VD category. Ns = *P* >.05, *= *P* ≤.05, and ****= *P* <0.00001. ^$^VD = Vitamin D, BMI = Body Mass Index, BAME = Black, Asian and Minority Ethnic.

### Age

Both age subgroups <50 and ≥50 years displayed similar differences in seropositivity with increasing VD (*Figure 2B*). There was only a significant difference in seropositivity between the severely deficient and insufficient VD levels in the <50 years subgroup (*P* = 0.03) (*Table 3*).

**Table 3.** P value table: comparisons of SARS-CoV-2 seropositivity within subgroups between VD category pairs.

|  | VD Category Comparison Pairs |  |  |  |  |  |
| --- | --- | --- | --- | --- | --- | --- |
|  | **1 & 2** | **1 & 3** | **1 & 4** | **2 & 3** | **2 & 4** | **3 & 4** |
| **Total Population** | **0.001** | **0.008** | **0.049** | 0.30 | 0.27 | 0.77 |
| **Male** | **0.006** | **0.01** | **0.004** | 0.62 | 0.74 | 0.36 |
| **Female** | **0.048** | 0.19 | 0.67 | 0.37 | 0.15 | 0.42 |
| **Age ≥50 (years)** | 0.48 | 0.23 | 0.50 | 1 | 1 | 0.81 |
| **Age <50 (years)** | 0.08 | **0.03** | 0.08 | 0.18 | 0.19 | 0.86 |
| **BAME** | 0.08 | **0.02** | 0.52 | 0.78 | 0.50 | 0.22 |
| **White** | 0.20 | 0.62 | 0.59 | 0.20 | 0.48 | 0.80 |
| **BMI <30 (Kg/m<sup>2</sup>)** | **0.01** | **0.03** | 0.07 | 0.35 | 0.48 | 1 |
| **BMI ≥30 (Kg/m<sup>2</sup>)** | 0.07 | 0.13 | 1 | 0.81 | 0.28 | 0.31 |
| **≥1 Comorbidity** | 0.08 | **0.045** | 0.30 | 1 | 0.43 | 0.28 |
| **0 Comorbidities** | **0.005** | 0.16 | 0.10 | 0.11 | 0.43 | 0.70 |

All comparisons were made with Fisher's exact test. Significant values ( $P<.05$ ) are **highlighted**. \*VD = Vitamin D, BMI = Body Mass Index, BAME = Black, Asian and Minority Ethnic.

### Ethnicity

Change in seropositivity rate with VD level was markedly different between the BAME and white ethnicity subgroups (*Figure 2C*). The BAME subgroup had a significant decline in seropositivity rate between severely deficient (78.6%) and insufficient (50.0%) positions; *P* = 0.02 (*Table 2* and *3*). The white subgroup presented a much slower decline in seropositivity rate and did not reach significance (*P* = 0.62) (*Table 3*). Both subgroups showed an increase in seropositivity rate with increasing VD from ~80-90 nmol/L, but seropositivity rate increased at a greater rate in the BAME subgroup.

### Sex

In both groups of men and women there was a significant decline in positivity between the severely deficient and deficient graph areas (*P* = 0.006 and *P* = 0.048 respectively) (*Table 3, Figure 2D*). Significant difference was found between the severe deficiency category and insufficiency category, and between the severe deficiency category and sufficiency category (*P* = 0.01 and *P* = 0.004 respectively) in the male subgroup (*Table 3*). Seropositivity rate was higher in women in the sufficiency category (61%) but did not reach significance (*P* = 0.13) (*Table 2* and *4*). Men reached a plateau in seropositivity rate at a lower VD level (~80 nmol/L) compared to women (~100 nmol/L).

### BMI

With regards to BMI, the change in seropositivity rate with VD level for both subgroups (≥30 Kg/m^2^ and <30 Kg/m^2^) was similar (*Figure 2E*). Both groups’ seropositivity rates declined until VD~80 nmol/L. A significant reduction in seropositivity rate occurred between both the severely deficient and the deficient category, and between the severely deficient and insufficient categories (*P* = 0.01 and *P* = 0.03 respectively) in the <30 Kg/m^2^BMI cohort (*Table 3*).

### Comorbidities

There were no differences in seropositivity rates between the comorbidity subgroups (0 or 1 or more) with vitamin D groups (*Figure 2F*). A significant reduction existed between the severely deficient and deficient areas in the 0 comorbidity group (*P* = 0.005) (*Table 3*). There was also significant difference between the severely deficient and insufficient categories in the 1+ comorbidities subgroup (*P* = 0.045) (*Table 3*). The largest difference in seropositivity rate existed within the insufficient graph area, with 44.1% in the 1+ comorbidities subgroup, and 60.2% in the 0 comorbidity subgroup (*P* = 0.052) (*Table 2* and *4*).

### Groups at risk of severe VD deficiency

Vitamin D deficiency is often related to age, sex, BMI, ethnicity, and co-morbidity so we assessed the impact of these on vitamin D levels and COVID-19 serology seropositivity.

### Age

There were no differences in the proportion of <50 year group in the severely deficient category in comparison to the ≥50 year group (*Figure 2B, Table 1*). Within the four VD categories, there was a higher proportion of <50 year individuals represented in all of them (ranging from 62.9-78.6%) (*Table 1*). In the severely deficient category, the ratio of ≥50 years: <50 years was 13:47 (i.e. 0.28:1) (*Table 1* and *4*). The ratio in the sufficient category was larger but did not meet statistical significance (*P* = 0.06), at 0.59:1 (*Table 1* and *5*). The largest difference existed between the sufficient category and the deficient category (0.27:1) (*Table 1*). This difference reached significance (*P* = 0.04) meaning an individual <50 years is 3.7 times more likely to be VD deficient than VD sufficient (*Table 1* and *5*).

### Ethnicity

Within the severely deficient category, there was a significantly larger proportion of the BAME group (40%) in the category in comparison to the white ethnic subgroup (6.6%); *p*-value <0.001 (*Table 1, Figure 2C*). The ratio of BAME: White in the severely deficient category (2.3:1) was significantly higher than the BAME: White ratios formed from each of the deficient, insufficient, and sufficient VD categories (0.26, 0.21 and 0.32:1 respectively; all *P* <0.001) (*Table 4, Figure 3C*). This indicates that BAME are more likely to be severely VD deficient than VD deficient.

**Table 4.** *P* value comparisons between VD Category pairs.

| Seroconversion |  | VD Category Comparison Pairs |  |  |  |  |  |
| --- | --- | --- | --- | --- | --- | --- | --- |
|  |  | 1 & 2 | 1 & 3 | 1 & 4 | 2 & 3 | 2 & 4 | 3 & 4 |
| **1** | Age (years) | 0.89 | 0.32 | 0.22 | 0.29 | 0.23 | 0.68 |
|  | Age ≥ 50 (years) | 0.80 | 0.52 | 0.14 | 0.83 | 0.34 | 0.40 |
|  | Sex | 0.18 | **0.04** | **0.01** | 0.66 | 0.17 | 0.32 |
|  | BMI (Kg/m<sup>2</sup>) | 0.99 | 0.32 | 0.28 | 0.33 | 0.34 | 0.97 |
|  | BMI ≥ 30 (Kg/m<sup>2</sup>) | 0.65 | 0.66 | 0.43 | 0.21 | 0.14 | 0.64 |
|  | Ethnicity | **< 0.00001** | **< 0.00001** | **0.0001** | 0.34 | 0.63 | 0.09 |
|  | Comorbidities | 0.50 | 0.85 | 0.50 | 0.24 | 0.11 | 0.55 |
| **0** | Age (years) | 0.63 | 0.07 | **0.006** | **0.03** | **0.001** | 0.08 |
|  | Age ≥ 50 (years) | 0.72 | 0.77 | 0.52 | 0.10 | 0.07 | 0.65 |
|  | Sex | 0.53 | 0.75 | 0.51 | 0.68 | 1 | 0.63 |
|  | BMI (Kg/m<sup>2</sup>) | 0.43 | 0.22 | 0.80 | 0.98 | 0.19 | 0.07 |
|  | BMI ≥ 30 (Kg/m<sup>2</sup>) | 0.56 | 1 | 0.21 | 0.32 | **0.02** | 0.11 |
|  | Ethnicity | **0.003** | **0.003** | **0.01** | 1 | 1 | 1 |
|  | Comorbidities | 0.54 | 0.28 | 1 | **0.01** | 0.24 | 0.39 |
| **Total** | Age (years) | 0.83 | 0.07 | **0.004** | **0.02** | **<0.001** | 0.11 |
|  | Age ≥ 50 (years) | 1 | 0.31 | 0.06 | 0.19 | **0.04** | 0.28 |
|  | Sex | 0.37 | 0.09 | 0.07 | 0.46 | 0.28 | 0.73 |
|  | BMI (Kg/m<sup>2</sup>) | 0.74 | 0.88 | 0.25 | 0.49 | 0.11 | 0.20 |
|  | BMI ≥ 30 (Kg/m<sup>2</sup>) | 0.38 | 0.86 | 0.12 | 0.19 | **0.01** | 0.08 |
|  | Ethnicity | **< 0.00001** | **< 0.00001** | **< 0.00001** | 0.62 | 0.58 | 0.27 |
|  | Comorbidities | 0.37 | 0.22 | 0.37 | **0.007** | **0.048** | 0.77 |

Comparisons were made within COVID-19 positive (1), COVID-19 negative (0), and the entire cohort.
Comparisons of age and BMI were made with Mann-Whitney U test, and binary variables for age, sex, BMI, ethnicity, and comorbidities used Fisher's exact test. Significant values (P<.05) are **highlighted**. \*VD = Vitamin D, BMI = Body Mass Index, BAME = Black, Asian and Minority Ethnic.

**Figure 3.**
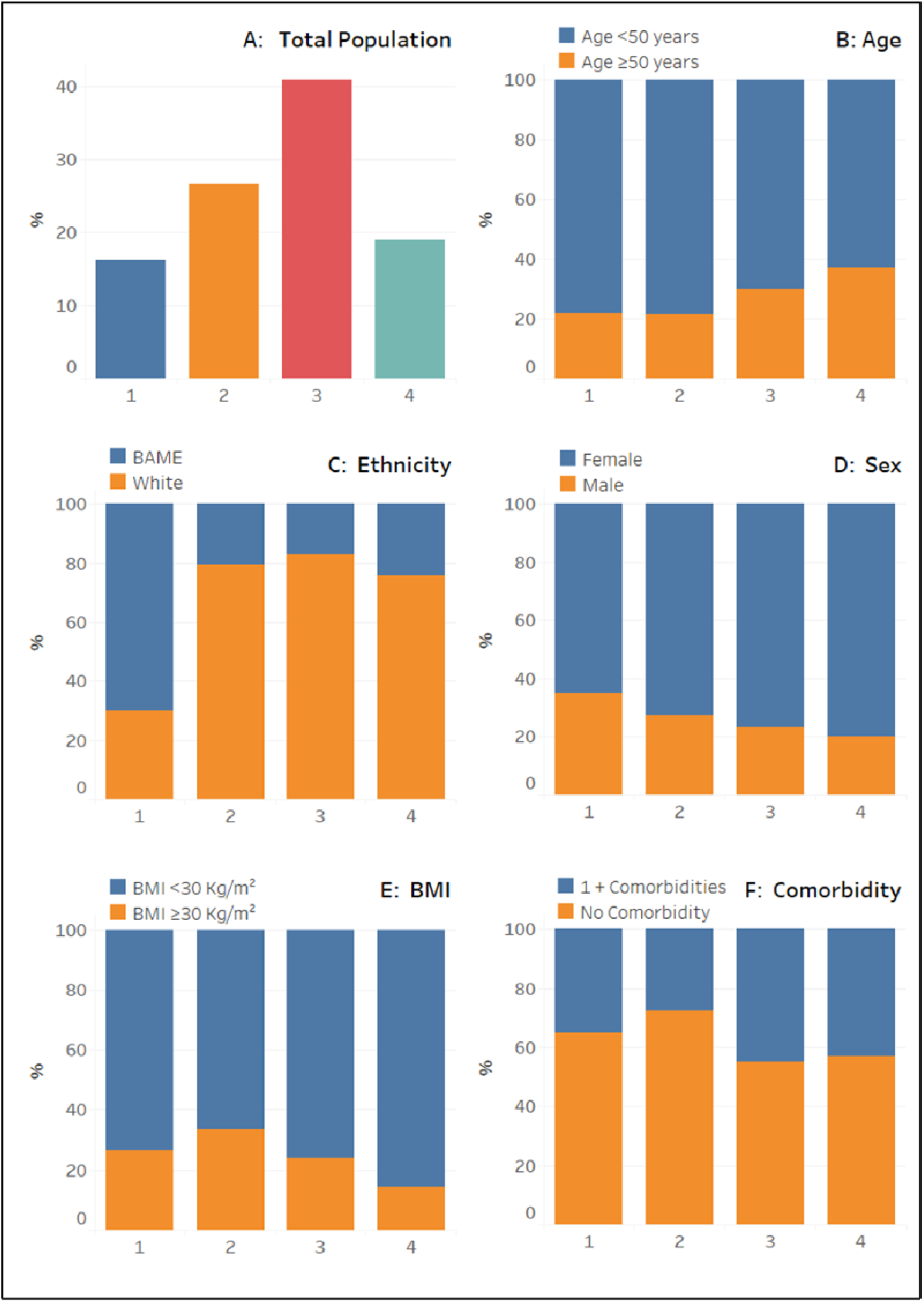
Relative proportions of paired subgroups within VD categories. Statistical comparisons in the proportions between VD categories is detailed in Table 4. VD category numbers: 1=Severe deficiency, 2=Deficiency, 3=Insufficiency, 4=Sufficiency. *VD = Vitamin D, BMI = Body Mass Index, BAME = Black, Asian and Minority Ethnic.

### Sex

There were no differences in the proportion of males compared to females in both severely VD deficient and VD deficient (*Figure 2D*). There were no significant differences existed between the proportions of any of the VD comparisons (*Figure 3D, Table 4*). The closest to significance (*P* = 0.07) existed between the severely deficient and sufficient group, with a ratio (Male: Female) of 0.54:1 and 0.25:1 respectively (*Table 1* and *5*).

### BMI

There were no significant differences in the proportion of the BMI ≥30 Kg/m^2^ subgroup in the severely VDD category *vs*. BMI <30 Kg/m^2^ subgroup (*Figure 2E*). A higher proportion of the BMI <30 Kg/m^2^ subgroup existed within all the VD categories (ranging between 66.3 – 85.7%) (*Table 1, Figure 3E*). Significant difference in the proportions was only found between VD categories of deficient and sufficient (*P* = 0.01) (*Table 4*). The ratio of BMI <30 Kg/m^2^ : ≥30 Kg/m^2^ was 1.94:1 and 6.14:1 respectively (*Table 1*).

### Comorbidity status

The proportion of the 0 comorbidity subgroup (which were VDD) was significantly higher (30.5%) compared to the equivalent in the 1+ comorbidities group (18.5%); *P* = 0.046 (*Figure 2F*). In both comorbidity subgroups, the proportions were higher in the deficiency category than in the severe deficiency category. A larger proportion of the 0 comorbidity group existed within each VD category compared to the 1+ comorbidity group (ranging from 55.0 – 72.4%) (*Table 1, Figure 3F*). The most equal proportions between the two comorbidity subgroups existed within the Insufficiency category (0.82:1) (*Table 1*). Significant difference was found between the insufficiency category and the deficiency category (0.38:1); *P* = 0.007 (*Table 1* and *5*). Significant difference was also found between the deficiency category and sufficiency category (*P* = 0.048) (*Table 4*).

## Discussion

In summary significant differences in SARS-CoV-2 seropositivity were found within the entire cohort, and several subgroups across the four VD categories. The sub-groups included both sex sub-groups; age >50 subgroup; BAME subgroup; BMI <30 Kg/m^2^ subgroup and both comorbidity subgroups. There were no significant differences between the seropositivity rates in the paired subgroups in any VD category. All curves displayed varying U-shaped curves, with ethnicity showing the most variation between subgroups: BAME subjects showed marked increase in seropositivity as VD level moved into the deficient areas, whereas the White cohort showed less variation in seropositivity across the VD continuum.

Seropositivity rate was significantly higher in the severely deficient category in comparison with any of the other VD categories within both the total population and the male subgroup. After a decline in seropositivity rate between a VD level of 30nmol/L and ~80nmol/L, an unexpected increase in seropositivity rate was apparent beyond VD ~80 nmol/L, resulting in a U-shaped curve that was reflected within all the subgroups.

A significantly larger proportion of the BAME population were severely VD deficient relative to the total white population. This finding was consistent across the total population, but also both the SARS-CoV-2 positive and negative BAME groups individually. These results add to the literature regarding individuals of darker skin are more likely to be VD deficiency because of a greater melanin content, reducing the availability of UVB rays for VD_3_ synthesis [7]. This evidence is also consistent with a recent study in the UK which found a significantly higher proportion of VD deficiency amongst new-borns within the BAME ethnic group [13]. Regardless of whether VDD is a cause or consequence (or both) of COVID-19, further investigation into VD supplementation in those of BAME ethnic group is warranted.

The U-shaped curves observed in this study challenges the current understanding of the relationship between VD and COVID-19, assumed to decline with increasing VD [14]. No published studies regarding COVID-19 have reproduced such results – i.e. those demonstrating increasing seropositivity at both ends of the VD spectrum. This U-shaped curve, however, has appeared in the wider literature regarding VD. For example, a large sample (n=24,094) study by Amrein et al. (2014) investigated the relationship between hospital admission VD and mortality [15]. Interestingly, after a decline in mortality with increasing VD, 90-day mortality rate began to increase beyond ~125 nmol/L levels, with an independent predictor of mortality beyond 150 nmol/L. In contrast, a review in 2016 identified all the studies (at the time) that investigated VD level against multiple outcomes within a U-shaped distribution: they concluded that the results were unlikely to be valid due to the lack of consideration of vulnerable individuals taking VD supplementation [16]. However, this study only had eight individuals on VD supplementation, from which only two had VD levels over 80 nmol/L-from a total of 51 subjects.

The only studies where the U-shaped curves were possibly significant were associated with allergy, due to a changing balance in the Th1/Th2 axis [16]. Due to U-shaped curves being identified in several studies against disease risk, further investigation is required to understand this phenomenon. One common explanation is whether the cause of increased disease risk passed a certain VD is due to the individual being on supplementation due to being clinically vulnerable already. However, our cohort were healthcare staff with few comorbidities. One possible mechanism of the U-shape curve could be due to induction of fibroblast Growth Factor-23 (FGF23) at higher levels of 25(OH)D (>100 nmol/L 25(OH)D and the consequent inhibition of 1-hydroxylase in immune cells [17].

## Limitations

There are a number of limitations to this study. The aggregation of multiple ethnicities into a singular ‘BAME’ subgroup provided a challenge, and due to the study population did not allow further sub-categorisation of ethnicity for which there may be further susceptibilities [6]. Another issue which is raised consistently in similar literature is seasonal variability in VD [18], however the participants were recruited within a tight timeframe in May, which should reduce effect of seasonal variation. Inclusion bias is another limitation raised consistently; individuals were recruited based on displayed symptoms of COVID-19 and isolation. This potentially increased the risk of selecting individuals that were more susceptible to COVID-19, regardless of their VD levels. Another limitation is that we have looked at seropositivity, rather than infection, though the used assay has a very high sensitivity for PCR proven disease. The amount of time the subjects may have been infected was not considered, and so VD levels *vs*. severity of disease (alongside confounding influences) will affect ability to interpret the results. Finally, the U-shaped curves were derived by grouping the samples into four VD bins, principally because there were limited numbers at each end of the VD spectrum.

## Conclusion

Our study has shown U-shaped relationship for COVID-19 seropositivity in UK healthcare workers. Further investigation is required to determine whether high VD levels can have a detrimental effect on COVID-19 susceptibility. Future randomised clinical trials of VD supplementation could potentially identify ‘optimal’ VD levels. This would allow for targeted therapeutic treatment for at-risk groups such as those within the BAME ethnic group.

## Data Availability

Data are available upon reasonable request. Proposals should be directed to the corresponding author.

## Declarations

### Ethics approval and consent to participate

All clinical data associated with the participants was pseudonymised and linked to the participant by a study ID number. The data was stored in an encrypted form in the secure University of Birmingham server, in accordance with ISO 27001/BS7799 information security guidance. The studies was approved by the London-Camden and Kings Cross Research Ethics Committee (20/HRA/1817).

### Authors’ contributions

DRT and AS conceptualised the study. AAF, SEF, CW, JED, AS, AGR and DRT contributed to data acquisition. WRM, STL, and AS analysed the data. All authors contributed to data interpretation. WRM, STL, DP, AS, and DRT drafted the manuscript. All authors contributed to the review and approval of the final copy of the manuscript.

## Acknowledgements

We thank the staff of University Hospitals Birmingham NHS Foundation Trust who kindly volunteered for this study. We would also like to thank the research staff of the Birmingham Wellcome NIHR Clinical Research Facility who undertook the staff facing assessments. We would like to thank colleagues at the Clinical Immunology Service for overseeing recruitment and sample processing.

